# Epidemiological profile and effectiveness of immediate postpartum contraception in Brazilian women

**DOI:** 10.1101/2022.08.12.22278711

**Authors:** Marcelo Luis Steiner, Júlia Lorenzini Mendes, Silvana Aparecida Giovanelli, Mônica Carneiro, Rodolfo Strufaldi, Mariliza Henrique da Silva

## Abstract

We analyzed the epidemiological profile of women who inserted copper intrauterine device (Cu-IUD), subdermal etonogestrel implant (ENG), tubal ligation (TL), depot medroxyprogesterone acetate (DMPA) or did not choose a contraceptive method (NCM) in the immediate postpartum. Also, we compared the contraceptive effectiveness of Cu-IUD and DPMA with non-MAC. Data from 20896 women were collected, of which 8183 (39%) opted for Cu-IUD, 559 (2.5%) DPMA, and 10989 (52.5%) for any method. When comparing these groups, women in the DPMA were younger (26.5±7.3, p<0.05), and NCM showed women with a lower number of pregnancies (2.2±1.3, p<0.05). Subjects in the TL group (4,6%) had the higher number of pregnancies (3,8±1.2, p<0.05), and ENG group, the highest number of miscarriages (1.6±1.3, p<0.05). Of those women who returned pregnant, 5.5% belonged to the DPMA group, 6% to the NCM group, and 2.3% to the Cu-IUD. Women who opted for Cu-IUD insertion were younger, had more pregnancies and vaginal delivery when compared to those who did not choose a method. Of those women who returned, the minority opted for Cu-IUD compared to those that opted for DPMA or no method.

## INTRODUCTION

Contraceptive methods play a central role in sexual and reproductive planning, enabling the couple to decide on their pregnancy planning freely and responsibly. [1] However, despite the ample supply of methods, about half of pregnancies worldwide are unplanned. [2]

Unplanned pregnancy is associated with a higher risk of perinatal complications, accounting for 60% of maternal deaths and 57% of child deaths. [3] Furthermore, the risks are more significant if these pregnancies occur within less than eighteen months, with a 61% low birth weight risk and 40% premature birth. [4]

The immediate postpartum period can be an opportune time to start a contraceptive method, as pregnant women are usually interested in preventing a new pregnancy and the hospital environment offers a facilitating situation for both doctor and patient. [5]

Long-acting reversible contraceptives (LARCs), such as intrauterine devices and hormonal implants, are good options for immediate postpartum contraception. [6] Besides having high satisfaction rates, continued use, and high efficacy, [8] the WHO has released them for use within the first 48 hours after delivery, provided the woman has no contraindications and chooses to leave the maternity hospital with a contraceptive method. [6]

A viable option of LARCs for use in the public health system in Brazil is the Copper Intrauterine Device (Cu-IUD) since they are available in the public health network, have high efficacy, have few side effects, and low cost. [1] The main concern of its insertion in the immediate postpartum period is expulsion, whose rates can vary from 0 to 13% [5, 6] and tend to be higher than in other elective insertion situations. [6, 7]

There is little information in the literature about the epidemiological profile of Brazilian women who accept or do not accept contraception in the immediate postpartum period and whether the method of choice is effective in the long run.

Aiming to contribute to a better general understanding of contraception in the immediate postpartum period, we assess the epidemiological profile of users according to the type of method chosen, and the rate of effectiveness in a municipal maternity hospital in São Bernardo do Campo.

## MATERIALS AND METHODS

### Methodology

A cross-sectional study at the University Hospital of São Bernardo do Campo (HMU-SBC) evaluated the electronic medical records of postpartum women enrolled from January 2016 to December 2020. The study was approved by the Hospital and ABC School of Medicine ethics committees.

We accessed all electronic medical records of postpartum women assisted at the HMU-SBC older than 18 years. Moreover, they were grouped as those who had inserted intrauterine cooper device intrapartum (Cu-IUD), underwent a tubal ligation during cesarean section (TL), had inserted a subdermal implant of etonogestrel (ENG), or had injected 150 mg of medroxyprogesterone acetate (DMPA). Those who did not choose contraception methods were considered “no contraception method” (NCM). Cu-IUD, ENG, and DMPA information were crosscheck with the hospital pharmacy’s medication dispensing control.

During hospital admission, the parturient needs to answer a standardized paper sheet containing questions about age, marital status, education degree, ethnicity, religious beliefs, and last pregnancies outcomes. In addition, cigarette, alcoholic beverages, and illicit drug use during the actual and previous pregnancy. Also, about utilization and type of contraceptive method at conception and if the pregnancy was planned, desired, or accepted. All these data and the type of delivery, information found elsewhere in the medical record, were collected.

The marital status was classified into two groups, married and stable union or single and divorced; ethnicity in Caucasian, African American, mixed (Caucasian with African American) or Asian; education degree in none, low, elementary, high school and postgraduation and religious beliefs in Catholic, Evangelical, Spiritism, Afro-Brazilian, other and no religion. Contraception use at the time of conception in “yes” or “no” and contraception type in oral or injectable contraceptive, intrauterine device, condom, or other. Cigarette, alcoholic beverages, illicit drug use, planned, desired, and accepted pregnancy were classified as “yes” or “no.”

Finally, we identified women who returned for second delivery during the study period and analyzed the contraceptive method chosen in the first hospitalization. Then the pregnancies interval and the sociodemographic characteristics were analyzed according to contraceptive method type.

### Statistical data analysis

Numerical variables were treated as mean and standard deviation, and qualitative variables as absolute numbers and percentages. The verification of the distribution of normality was performed using the Shapiro-Wilk test. The comparison between groups with normal distribution was performed using the one-way ANOVA test, those without normal distribution Kruskal-Wallis test, and for qualitative variables Chi-square test. SPSS version 2019 software was used. Statistical tests were considered significant if the p-value was less than 5%.

### Patient and public involvement

This study has not a direct patient and public involvement. Eletronic medical records of postpartum women enrolled from January 2016 to December 2020 were collected. This group was not recruited and did not received any intervention, just a retrospective analyses of its characteristics.

## Results

As shown in table 1, data from 20,896 women were collected, of which 8,183 (39%) had Cu-IUD inserted; 961 (4.5%) performed tubal ligation; 559 (3%) chose DPMA; 204 (1%) ENG and 10,989 (52.5%) chose not to use contraception at the time of hospital discharge. When comparing the groups, those from the DPMA and ENG were younger, and those from the TL group were older (p<0.05). The NCM group had the lowest number of pregnancies (2.2±1.3), and the TL group had the highest (3.8±1.2) when compared to the other groups (p<0.05). Women referred for ENG had the highest number of vaginal births (2.4±1.8), and those who underwent TL had the highest number of cesarean sections (2±0.6) in comparison with the other groups (p<0.05).

**Table 1.** Epidemiological characteristics of the population.

|  |  | IUD<br>(N=8183) | Tubal<br>Ligation<br>(N=961) | DMPA<br>(N=559) | Implant<br>(N=204) | WC (N=10989) | p |
| --- | --- | --- | --- | --- | --- | --- | --- |
|  |  | m±sd | m±sd | m±sd | m±sd | m±sd |  |
| Age |  | 28±6.7<sup>a</sup> | 35 ± 5.4<sup>b</sup> | 26.5±7.3<sup>c</sup> | 27±8<sup>d</sup> | 29±7<sup>e</sup> | **<0.01** |
| Number of pregnancies |  | 2.3±1.4<sup>f</sup> | 3.8±1.2<sup>b</sup> | 2.4±1.6<sup>f</sup> | 3±2.2<sup>d</sup> | 2.2±1.3<sup>e</sup> | **<0.01** |
| Normal Birth |  | 1.8±1.2<sup>f</sup> | 2±1.2 | 2±1.3 | 2.4±1.8<sup>g</sup> | 1.7±1<sup>e</sup> | **<0.01** |
| Cesarean Section |  | 1.2±0.5 | 2±0.6<sup>b</sup> | 1.2±0.5 | 1.1±0.4 | 1.2±1 | **<0.01** |
| Number of Abortions |  | 1.2±0.6<sup>h</sup> | 1.4±0.7 | 1.35±0.8 | 1.6±1.3 | 1.3±0.7 | **<0.01** |
|  |  | N(%) | N(%) | N(%) | N(%) | N(%) |  |
| Type of Delivery | Normal | 4940<br>(62.7) | 15 (0.1) | 358 (75) | 110 (70) | 5570 (66.8) | **0.01** |
|  | Cesarean Section | 2929<br>(37.2) | 892 (98) | 114 (24.1) | 46 (29.5) | 2763 (33.1) |  |
| Ethnicity | Caucasian | 2795 (43) | 412 (42.8) | 213 (38.1) | 64 (31.3) | 4756 (43.3) | **0.01** |
|  | Mixed | 3273<br>(50.3) | 485 (50.4) | 304 (54.3) | 111<br>(54.4) | 5392 (49.1) |  |
|  | Afro-American | 322 (4.9) | 51 (5.3) | 28 (5) | 13 (6.3) | 551 (5) |  |
|  | Asian | 9 (0.1) | 2 (0.2) | 2 (0.3) | 1 (0.5) | 31 (0.2) |  |
| Marital Status | Married - Stable Union | 4616 (70) | 784 (81) | 342 (61.1) | 98 (48) | 7753 (70.6) | **0.01** |
|  | Single – Divorced | 1886 (29) | 177 (18.4) | 214 (38.4) | 106 (52) | 3215 (30) |  |
| Education Degree | None | 5 (0.07) | 5 (0.5) | 3 (0.5) | 5 (2.4) | 15 (0.1) | **0.01** |
|  | Low | 204 (3.13) | 43 (4.4) | 14 (2.5) | 25 (12.2) | 364 (3.3) |  |
|  | Elementary | 1210<br>(18.6) | 200 (20.8) | 125 (22.3) | 91 (44.6) | 1908 (17.3) |  |
|  | High School | 4062<br>(62.4) | 564 (58.6) | 377 (67.4) | 67 (32.8) | 6715 (61.1) |  |
|  | Postgraduation | 841 (13) | 123 (12.7) | 34 (6) | 7 (3.4) | 1594 (14.5) |  |
| Religion | Catholic | 1948 (24) | 332 (34.5) | 120 (21.5) | 40 (19.5) | 3303 (30) | **0.01** |
|  | Evangelical | 2865 (35) | 455 (47) | 264 (47) | 77 (38) | 4720 (43) |  |
|  | Spiritism | 98 (1) | 21 (2) | 7 (1.5) | 2 (1) | 160 (1.5) |  |
|  | Afro-braziliann | 78 (1) | 7 (1) | 5 (1) | 3 (1.5) | 110 (1) |  |
|  | No Religion | 631 (8) | 55 (6) | 80 (14) | 40 (19.5) | 1034 (9.5) |  |
|  | Other Religions | 2563 (31) | 91 (9.5) | 83 (15) | 42 (20) | 1648 (15) |  |
| Contraception | Yes | 2182 (34) | 368 (39) | 173 (31.3) | 38 (19.3) | 3419 (31.8) | **0.01** |
|  | No | 4218 (66) | 573 (60) | 379 (68.6) | 159 (80.7) | 7307 (68.1) |  |
| Type of Contraception | Oral Contraception | 1478 (22.7) | 220 (0.5) | 111 (19.8) | 17 (8.3) | 2231 (20.3) | **0.01** |
|  | Injectable contraceptive | 210 (3.2) | 40 (4.1) | 21 (3.7) | 8 (3.9) | 305 (2.7) |  |
|  | IUD | 23 (0.3) | 13 (1.3) | 10 (1.7) | 1 (0.4) | 71 (0.6) |  |
|  | Condom | 277 (4.2) | 57 (6) | 22 (3.9) | 8 (3.9) | 525 (4.7) |  |
|  | Other | 4520 (69.4) | 630 (65.6) | 395 (70.6) | 170 (83.3) | 7860 (71.4) |  |
| Planned Pregnancy | Yes | 1804 (28.1) | 216 (22.6) | 151 (27.4) | 28 (14) | 3722 (34.7) | **0.01** |
|  | No | 4607 (71.8) | 736 (77.3) | 400 (72.5) | 172 (86) | 7006 (65.3) |  |
| Desired Pregnancy | Yes | 4762 (75) | 658 (70) | 405 (74) | 101 (52) | 8296 (78) | **0.01** |
|  | No | 1613 (25) | 281 (30) | 143 (26) | 94 (48) | 2332 (22) |  |
| Accepted Pregnancy | Yes | 6456 (91.5) | 918 (98) | 524 (96) | 162 (84.8) | 10280 (97.3) | **0.01** |
|  | No | 598 (8.4) | 18 (2) | 22 (4) | 29 (15.2) | 283 (2.7) |  |
| Number of pregnancies | Primiparous | 2591 (31.7) | 9 (0.9) | 195 (35) | 63 (31) | 3994 (36.6) | **0.01** |
|  | Multiparous | 5570 (68.2) | 951 (99) | 363 (65) | 140 (69) | 6906 (63.3) |  |
| Tobacco Use | Yes | 754 (11.6) | 141 (14.7) | 90 (16.1) | 118 (57.8) | 1115 (10.1) | **0.01** |
|  | No | 5751 (88.4) | 820 (85.3) | 469 (83.9) | 86 (42.1) | 9860 (89.8) |  |
| Alcohol Use | Yes | 243 (3.7) | 29 (3) | 34 (6) | 64 (31.3) | 384 (3.5) | **0.01** |
|  | No | 6262 (96.2) | 932 (97) | 525 (94) | 140 (68.7) | 10591 (96.5) |  |
| Drug Use | Yes | 155 (2.4) | 22 (2.2) | 29 (5.1) | 103 (50.4) | 251 (2.3) | **0.01** |
|  | No | 6350 | 939 (97.7) | 530 (94.8) | 101 | 10724 (97.7) |  |

|  | (97.6) | (49.5) |
| --- | --- | --- |
| a- Bonferroni Test $p < 0.05$ LT Group versus other groups | | |
| b- Bonferroni Test $p < 0.05$ DPMA Group versus Cu-IUD, LT and Absent Groups | | |
| c- Bonferroni Test $p < 0.05$ IMP Group versus LT and Absent Groups | | |
| d- Bonferroni Test $p < 0.05$ Absent Group versus other groups | | |
| e- Bonferroni Test $p < 0.05$ Cu-IUD and DPMA Group versus LT, IMP and Absent Groups | | |
| f- Bonferroni Test $p < 0.05$ IMP Group versus other groups | | |
| g- Bonferroni Test $p < 0.05$ Cu-IUD Group versus LT and IMP Groups | | |

Of those women from the ENG group, only 48% reported having a stable family relationship or being married, while of those from TL, this number reached 81%. All groups, except for ENG, had more than 55% of women with high school completed. Women in the Cu-IUD were less catholic and protestant and showed a higher number of women with non-discriminated religions than the other groups. Except for the group that inserted ENG, the groups were similar concerning previous contraception methods use, pregnancy planning or not, smoking, alcohol consumption, and drug use.

As shown in table 2, 877 women returned to the hospital for a second birth in the period analyzed. Of these, 654 (74.5%) had not opted for contraception, 31 (3.5%) for DPMA, and 189 (21.5%) had inserted Cu-IUD. Two women had undergone tubal ligation, and only one had ENG. Considering the total 20,896 women evaluated, only 2.3% from the Cu-IUD group returned, while from the DMPA group, it was 5.5% and WC 6%. The average return time was shorter for those who opted for DMPA than the other groups. There was a higher percentage of evangelical women in the DMPA group and a lower percentage of programmed pregnancy in the Cu-IUD group.

**Table 2.** Personal characteristics, return time and pregnancy schedule of women who returned pregnant according to the choice of method in the immediate postpartum period (IUD, DMPA or without method)

|  |  | Cu-IUD<br>n=189 | DPMA<br>N=31 | WC N=654 | p |
| --- | --- | --- | --- | --- | --- |
| | | m ± sd | m ± sd | m ± sd | |
| Age | | 27.2±5.7 | 28.5±7 | 28±6.1 | 0.266 |
| Δt. Pregnancy (days) | | 697.1±252 | 591.5±222 | 692.1±252 | 0.087 |
|  |  | n% | n% | n% |  |
| ETHNICITY | Caucasian | 87 (46.5) | 10 (32) | 247 (38) | 0.072 |
|  | Non-Caucasian | 100 (53.5) | 21 (68) | 405 (62) |  |
| MARITAL STATUS | Married - Stable Union | 133 (70.5) | 21 (68) | 468 (71.5) | 0.867 |
|  | Single – Divorced | 56 (29.5) | 10 (32) | 186 (28.5) |  |
| EDUCATION | Low Education | 13 (7) | 4 (13.5) | 33 (5.5) | 0.094 |
|  | Elementary (seconda half) - High School | 157 (87) | 24 (80) | 520 (84) |  |
|  | Higher Education and Postgraduate Studies | 10 (6) | 2 (6.5) | 66 (10.5) |  |
| RELIGION | No Religion | 35 (19.5) | 7 (26) | 113 (19) | 0.005 |
|  | Evangelical | 90 (50) | 16 (59.5) | 279 (47) |  |
|  | Catholic | 52 (29) | 2 (7.5) | 180 (30.5) |  |
|  | Afro-Brazilian | 2 (1) | 0 (0) | 6 (1) |  |
|  | Spiritism | 1 (0.5) | 0 (0) | 9 (1.5) |  |
|  | Other | 0 (0) | 2 (7.5) | 4 (1) |  |
| PLANNED | Yes | 27 (14.5) | 10 (32.5) | 176 (27) | 0.001 |
| PREGNANCY | No | 162 (85.5) | 21 (67.5) | 478 (73) |  |

## DISCUSSION

In comparing the different contraception types at the early postpartum period, those who opted for the immediate insertion of a Cu-IUD were younger, prone to have a higher number of births and previous pregnancy than the group that chose not to have any method. Overall, the clinical profile between these two groups was very similar. Even those factors that showed a significant difference in clinical practice are subtle. Thus, the authors consider no specific profile of women candidates for immediate postpartum Cu-IUD insertion, and it should be offered for all women.

The epidemiological profile found in this study differs from that reported in a follow-up study of European women who had an intrauterine device (EURAS-IUD). At the time of insertion, the mean age of European women was 25±3.1, and the mean number of live births was 1.6; while in the population of this study, the mean was 28±6.7, and the number of previous pregnancies was 2.3±1.4. It should be noted that the European follow-up study evaluated women who had the IUD inserted at any time and not just postpartum. Also, we must consider whether the offer of this method is occurring at a late age in the city of São Bernardo do Campo, increasing the chance of unplanned pregnancy.

The occurrence of unplanned pregnancy was 71% and 65% in Cu-IUD and WC groups, respectively. It is noteworthy that, in both groups, the percentage of women using some contraceptive methods was slightly higher than 30%. The oral hormonal contraceptive was the most frequent in both groups (close to 20%). Considering many women without contraceptive methods and those who use low-effectiveness methods, we can assume that the study population does not have access or adequate health equipment for reproductive planning.

In this context, the possibility of providing counseling and establishing the use of effective methods makes prenatal care and hospitalization for childbirth care a window of opportunity for reproductive planning.

Of those women who underwent tubal ligation or subcutaneous hormonal implantation on admission, less than 0.5% returned to the hospital with a second pregnancy in the period evaluated, confirming ENG with a highly effective long-term reversible method. The return rate of pregnant women among those who inserted Cu-IUD was 2.3%, a number consistent with the literature and lower than that presented WC group, which had a return rate of 6%. The group that underwent DMPA had a return of 5.5%, demonstrating lower efficacy, with a rate close to WC group. This finding supports the argument that this method should not be considered a long-term method, as it requires a user action every three months.

Regardless of the contraception type group, the minority of returning women programmed the pregnancy. It is noteworthy that the women who opted for DPMA in the first hospitalization had a shorter interval between births than the other groups analyzed, which had similar intervals between them. The explanation for the shorter interval between pregnancies for the DPMA group is unclear. The possibility of amenorrhea associated with an injection performed in the hospital can give a feeling of security and decrease adherence to the method. Indeed, considering the success rate and the interval between pregnancies, the indication of this method should be put into perspective for this immediate postpartum population.

The women in this study who inserted hormonal implants have specific characteristics, but different from those found in the study by Eggebroten JL et al, they have a high average number of pregnancies and miscarriages; a higher percentage of unplanned pregnancy and most were single marital status, smokers, alcohol and drug users. The reason is that this method is only available in the hospital for women considered to be in a socially vulnerable situation.

Likewise, the group that underwent tubal sterilization has specific characteristics for being included in a protocol of legal criteria: more than two pregnancies or more than 25 years. Thus, it is expected that women who underwent TL are older and have more pregnancies than other groups.

The study has limitations. The return rate may be underestimated as it is impossible to follow up with all the women or even guarantee that all who became pregnant returned to the hospital. Also, the reliability of the data depends on correctly filling out the medical records, and the retrospective methodology does not allow for an assessment of the continuity and satisfaction of the methods used. On the other hand, the number of women, the different types of methods evaluated with data collected over five years allow for an adequate understanding and insights into reproductive planning in the immediate childbirth context.

According to our results and excluding women in vulnerable situations, there is no specific profile of women candidates for long-term contraceptive methods. The insertion of Cu-IUD and ENG in the postpartum period reduced the chance of returning with a new unplanned pregnancy, with a performance superior to DPMA.

Thus, prenatal care and immediate postpartum should be considered moments of opportunity for reproductive planning actions, especially with the guidance and performance of LARC.

## Data Availability

All data produced in the present work are contained in the manuscript

## ACKNOWLEDGMENTS

Everyone involved in the study. Hospital Municipal Universitário de São Bernardo for the partnership and encouragement of research.

